# Identifying Message “Recipes” for High Perceived Effectiveness: Cognitive, Social, and Emotional Profiles of Vaping Public Education Messages Among Young Adults Who Vape

**DOI:** 10.64898/2026.10.04.26364669

**Authors:** Jiaying Liu, Qiyao Peng, Musa Malik, Yidi Wang, Emily Norton, Tianlai Ye, Colleen Markey, Allison Worsdale, Lawrence H. Sweet

## Abstract

**Introduction:** Vaping public education campaigns are a central prevention strategy for young adults (YAs), but message effectiveness varies widely and formative testing typically evaluates one message attribute at a time. This study examined how four perceptual dimensions—cognitive engagement, social relevance, emotional arousal, and emotional valence—relate to perceived message effectiveness (PME), and whether multiple combinations of these dimensions correspond to similarly high PME.

**Methods:** In a cross-sectional online message-testing study conducted from March to April 2025, 540 U.S. YAs who vape each evaluated 15 messages randomly sampled from a corpus of 229 text-image vaping public education messages, yielding 8,100 message evaluations. Cross-classified multilevel models estimated associations between each perceptual dimension and PME. An Explainable Boosting Machine probed nonlinear associations, and *k*-means clustering of high-predicted-PME perceptual configurations identified distinct profiles. Analyses were conducted in 2026.

**Results:** Cognitive engagement (*b* = 0.20, 95% CI [0.18, 0.22]) and social relevance (*b* = 0.15, 95% CI [0.13, 0.17]) showed the strongest associations with PME, followed by emotional arousal (*b* = 0.06, 95% CI [0.04, 0.08]); emotional valence showed no linear association (*b* = 0.00, 95% CI [−0.02, 0.02]). The Explainable Boosting Machine revealed a U-shaped valence pattern, with higher predicted PME at both the negative and positive ends of the scale. Clustering yielded five distinct perceptual profiles with similarly high predicted PME (4.10–4.25).

**Conclusions:** Multiple perceptual configurations correspond to similarly high PME, supporting a configurational approach to formative message testing rather than a single optimal message formula.

## Introduction

Nicotine vaping remains common among U.S. young adults (YAs). In 2025, 22.3% of YAs aged 18-25 years reported past-month nicotine vaping, the highest prevalence of any age group in the National Survey on Drug Use and Health.^1^ Although e-cigarettes may provide harm-reduction benefits for adults who smoke combustible cigarettes when used as a complete substitute, the public health implications differ for YAs, among whom vaping often occurs outside the context of cigarette smoking or cessation.^2^ In national data, most current YA e-cigarette users had never smoked cigarettes, underscoring the importance of preventing nicotine dependence and continued vaping in this population.^3^ Public education campaigns, are an important strategy for communicating the health risks of vaping and discouraging use among youth and YAs.^4^ Recent systematic reviews and meta-analyses indicate that vaping-related public education messages can increase risk perceptions and other cognitive outcomes and may reduce susceptibility to vaping, although effects vary across messages and outcomes.^5–7^

A central challenge in campaign development is therefore not simply whether prevention messages can work, but which messages are most promising for a particular audience. Formative message testing is routinely used to compare candidate messages before large-scale dissemination, with target-audience evaluations informing message selection and refinement. Recent FDA research further demonstrates the value of quantitative message evaluation during campaign development, including at the preproduction stage.^9^ One commonly used outcome is perceived message effectiveness (PME), which captures recipients’ evaluations of a message’s persuasive potential.^10–12^ Although PME is not equivalent to actual behavioral effectiveness, longitudinal and experimental evidence indicates that PME can anticipate subsequent vaping-related outcomes and is useful for distinguishing more versus less promising messages.^12–14^

Existing message-testing research has identified specific content and execution features associated with stronger audience responses. For example, harm- and addiction-related content, graphic or concrete depictions, and other design features can influence perceived effectiveness, whereas some flavor, vernacular, or social themes may perform less consistently.^7,15^ Recent work has extended this approach by systematically cataloging objective features of hundreds of vaping-prevention advertisements, including message themes, structural characteristics, product depictions, and audience targeting.^8^ These objective inventories provide an important foundation for campaign design, but observable message content does not fully describe how a message is experienced by its intended audience. The same objective feature may evoke different degrees of thought, social relevance, or emotional response, and these audience perceptions may jointly shape message evaluation.

Recent evidence supports considering these psychological responses together. In a randomized evaluation of FDA’s *The Real Cost* vaping-prevention public service announcements (PSAs), effects on vaping susceptibility were partly associated with cognitive elaboration, negative affect, and injunctive norms, alongside changes in risk beliefs and attitudes.^16^ Other recent studies similarly show that perceived relevance, emotional responses, and PME vary as a function of message content and framing among young people exposed to vaping prevention communications.^7,17,18^ These findings suggest that formative message testing may benefit from moving beyond isolated message attributes toward a more integrated characterization of how target audiences cognitively, socially, and emotionally experience messages.

Guided by this literature, in the present study we focused on four perceptual dimensions: cognitive engagement, social relevance, emotional arousal, and emotional valence. These dimensions capture complementary cognitive, social, and affective aspects of message experience that have been linked to persuasive processing and responses to tobacco-related messages. Cognitive engagement reflects the extent to which a message prompts effortful, elaborative processing, as emphasized in dual-process models of persuasion.^19,20^ Social relevance reflects the extent to which a message evokes interpersonal or social consequences, consistent with evidence that social and normative considerations can shape responses to tobacco-related messages, particularly among adolescents and YAs.^16^ Emotional arousal and valence distinguish the intensity and direction of affective responses, respectively, two well-established dimensions for characterizing emotional experience.^21^ Recent vaping-message research further suggests that cognitive elaboration, affective responses, and social or normative processes can contribute to message effects.^7,16^ These four dimensions are not intended to capture all aspects of message experience, but rather provide a parsimonious set of complementary perceptual characteristics that can be assessed consistently across a large and heterogeneous message set.

Building on this approach, the present study seeks to identify message “recipes,” or combinations of perceptual dimensions associated with high PME. Multiple young adults who vape independently evaluated each message, and their ratings were aggregated to generate message-level estimates of cognitive engagement, social relevance, emotional arousal, and emotional valence. We first examine the associations between each perceptual dimension and PME. We then use interpretable machine learning to identify nonlinear patterns and combinations of perceptual dimensions associated with high PME, and to derive distinct message “recipes” from these high-performing profiles. The study addresses two questions:

*RQ1*. Which cognitive, social, and emotional perceptual dimensions are most strongly associated with PME, and do these associations exhibit linear or nonlinear patterns?

*RQ2.* Which combinations of these perceptual dimensions characterize message “recipes” associated with high predicted PME?

## Methods

### Design and Participants

We conducted a cross-sectional online message-testing study from March to April 2025 with 540 U.S. YAs aged 18-29 years (*M*age = 24.4, *SD* = 3.1; 48% female) who reported vaping at least weekly during the past 30 days (*M* = 18.9, *SD* = 10.4). Participants were recruited through CloudResearch’s Managed Prime panel using prespecified quotas for sex assigned at birth, race, and Hispanic ethnicity intended to approximate U.S. Census demographic distributions among YAs.^22^ The final sample was predominantly White (71.5%), with additional representation from Black or African American (16.7%), Asian (4.3%), American Indian or Alaska Native (4.4%), and other racial groups. Approximately 25% identified as Hispanic or Latino/a/x. After providing consent, each participant viewed 15 messages randomly sampled without replacement from a set of 229 text-image vaping public education messages and completed perceptual ratings and PME after each exposure. This design yielded 8,100 message evaluations in total. The recruitment target was selected to provide approximately 25 independent evaluations per message, consistent with prior message-testing guidance.^23^ Because messages were randomly sampled for each participant, the number of evaluations per message varied (*M* = 35.4, *SD* = 6.1, range = 20-55). No data were missing on the perceptual dimensions or PME items.

### Stimuli

The stimulus corpus consisted of 229 static text-image vaping public education messages assembled to capture a broad range of message content and persuasive approaches relevant to YAs. Messages were drawn from three sources: (1) existing public education materials, including messages collected or adapted from well-known youth- and YA-focused tobacco and vaping campaigns and organizations such as *The Real Cost*, *truth initiative*, and *FlavorsHookKids*; (2) targeted modifications of online images and accompanying text; and (3) newly developed messages created through collaborative creative work with research team members and graphic designers. Both the modified and newly developed messages were guided by themes identified through prior formative research with YAs who vape,^24^ such as appearance-related health consequences, autonomy, family impact, and social impressions. These themes were also broadly consistent with prior research identifying promising targets for vaping prevention communication among youth and YAs.^25^ Together, these sources yielded a heterogeneous message corpus spanning cognitive, emotional, and social persuasive approaches, while allowing individual messages to incorporate elements from more than one approach. The full stimulus corpus is not publicly redistributed because some messages incorporate third-party campaign or online-source materials that may be subject to copyright restrictions. Example messages are shown in Table 1.

**Table 1.**
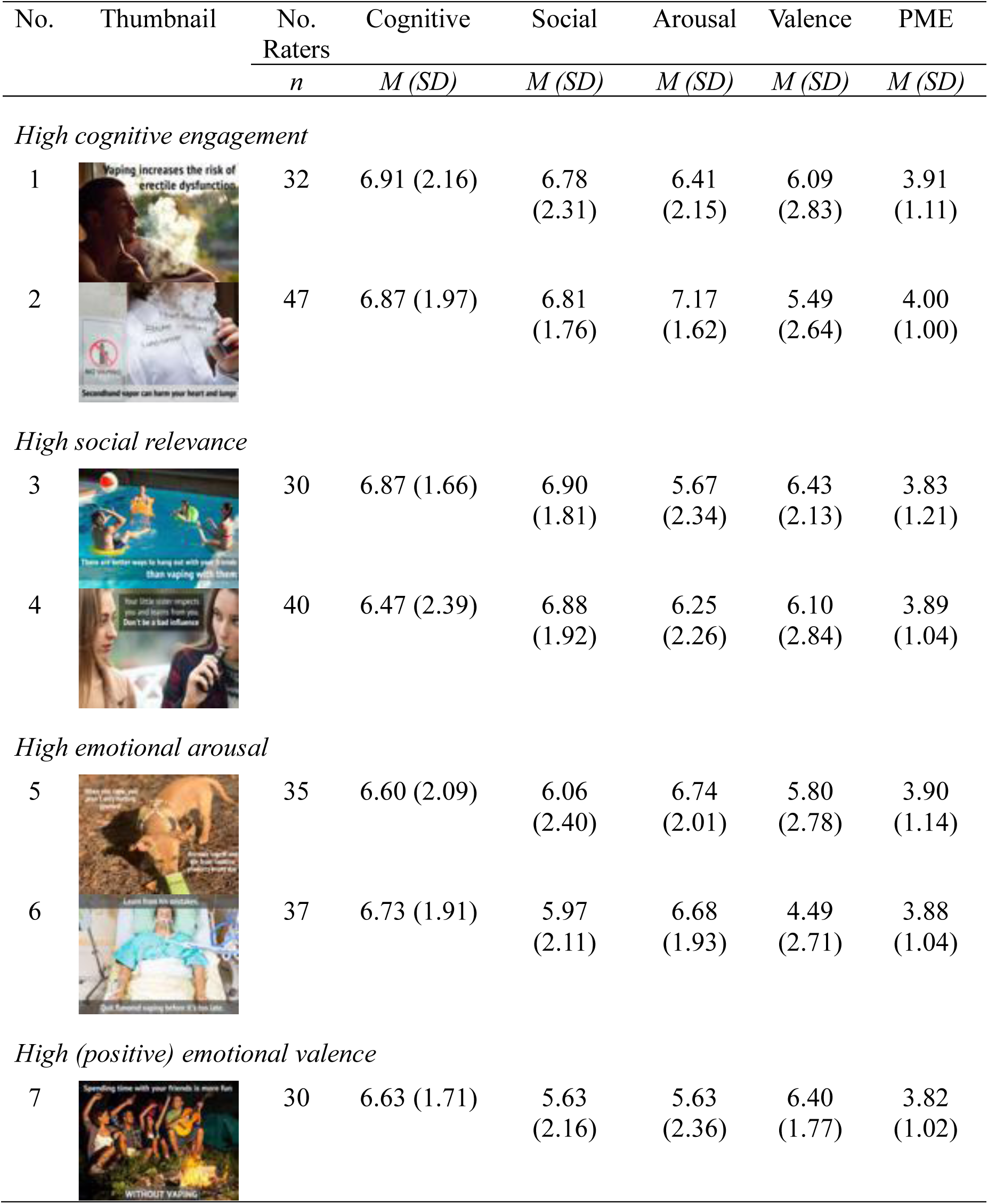
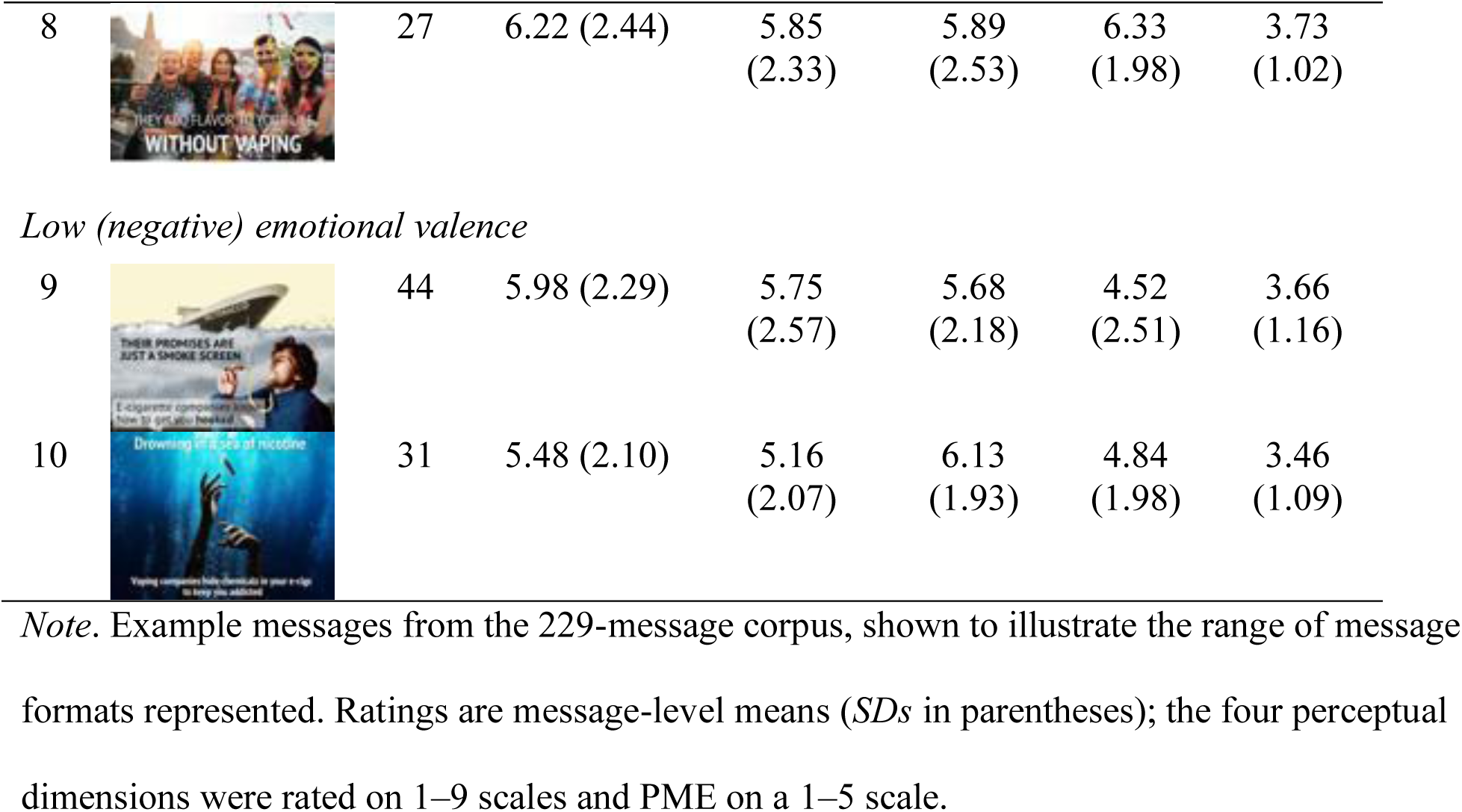
Illustrative Vaping Public Education Messages From the 229-Message Corpus.

| No. | Thumbnail | No.<br>Raters | Cognitive | Social | Arousal | Valence | PME |
| --- | --- | --- | --- | --- | --- | --- | --- |
|  |  | *n* | *M (SD)* | *M (SD)* | *M (SD)* | *M (SD)* | *M (SD)* |
| High cognitive engagement |  |  |  |  |  |  |  |
| 1 |  | 32 | 6.91 (2.16) | 6.78 (2.31) | 6.41 (2.15) | 6.09 (2.83) | 3.91 (1.11) |
| 2 |  | 47 | 6.87 (1.97) | 6.81 (1.76) | 7.17 (1.62) | 5.49 (2.64) | 4.00 (1.00) |
| High social relevance |  |  |  |  |  |  |  |
| 3 |  | 30 | 6.87 (1.66) | 6.90 (1.81) | 5.67 (2.34) | 6.43 (2.13) | 3.83 (1.21) |
| 4 |  | 40 | 6.47 (2.39) | 6.88 (1.92) | 6.25 (2.26) | 6.10 (2.84) | 3.89 (1.04) |
| High emotional arousal |  |  |  |  |  |  |  |
| 5 |  | 35 | 6.60 (2.09) | 6.06 (2.40) | 6.74 (2.01) | 5.80 (2.78) | 3.90 (1.14) |
| 6 |  | 37 | 6.73 (1.91) | 5.97 (2.11) | 6.68 (1.93) | 4.49 (2.71) | 3.88 (1.04) |
| High (positive) emotional valence |  |  |  |  |  |  |  |
| 7 |  | 30 | 6.63 (1.71) | 5.63 (2.16) | 5.63 (2.36) | 6.40 (1.77) | 3.82 (1.02) |
|  |  | *n* | *M (SD)* | *M (SD)* | *M (SD)* | *M (SD)* | *M (SD)* |
| 8 |  | 27 | 6.22 (2.44) | 5.85<br>(2.33) | 5.89<br>(2.53) | 6.33<br>(1.98) | 3.73<br>(1.02) |
| *Low (negative) emotional valence* |  |  |  |  |  |  |  |
| 9 |  | 44 | 5.98 (2.29) | 5.75<br>(2.57) | 5.68<br>(2.18) | 4.52<br>(2.51) | 3.66<br>(1.16) |
| 10 |  | 31 | 5.48 (2.10) | 5.16<br>(2.07) | 6.13<br>(1.93) | 4.84<br>(1.98) | 3.46<br>(1.09) |

*Note.* Example messages from the 229-message corpus, shown to illustrate the range of message formats represented. Ratings are message-level means (*SDs* in parentheses); the four perceptual dimensions were rated on 1–9 scales and PME on a 1–5 scale.

### Measures

#### Perceived message effectiveness

PME was assessed using a brief three-item message-perception measure adapted from prior PME research.^26,27^ Prior message-testing studies have included *convincing*, *effective*, and *believable* within multi-item assessments of PME,^26^ and factor-analytic work has shown that these closely related message-evaluation items load on a common PME dimension.^27^ Consistent with Noar and colleagues’ distinction between message perceptions and effects perceptions,^10^ the present measure focused on evaluations of the message itself rather than anticipated effects on recipients. Because each participant evaluated 15 messages, a brief measure was selected to reduce repetitive respondent burden while retaining core indicators of perceived persuasive quality. After each message exposure, participants rated their agreement that the message was *convincing*, *effective*, and *believable* on 5-point scales ranging from 1 (*strongly disagree*) to 5 (*strongly agree*). The three items were averaged to form an exposure-level PME score (Cronbach *α* = .81), with higher scores indicating greater PME. Message-level PME was calculated by averaging exposure-level PME scores across participants who evaluated each message.

#### Perceptual dimensions

Each message was evaluated on four perceptual dimensions: cognitive engagement, social relevance, emotional arousal, and emotional valence. Emotional arousal and valence were assessed using 9-point pictorial scales adapted from the Self-Assessment Manikin (SAM).^21^ Emotional valence ranged from 1 (*most unpleasant*) to 9 (*most pleasant*), with 5 representing neutral affect. Emotional arousal ranged from 1 (*most calm*) to 9 (*most stimulated*), with 5 representing a neutral or usual level of arousal/activation.

Cognitive engagement and social relevance were assessed using study-specific 9-point scales with a parallel pictorial response format, ranging from 1 (*not at all*) to 9 (*extremely*). Cognitive engagement assessed how thought-provoking the message was, with thought-provoking defined as prompting participants to reflect on their knowledge about vaping and its consequences. Social relevance assessed the extent to which the message brought to mind potential positive or negative social consequences of vaping. Higher scores indicated greater cognitive engagement and social relevance, respectively. Message-level scores for each dimension were computed by averaging participant ratings across all participants who evaluated a given message. Analytic transformations of these scores are described below.

#### Participant characteristics

Participants reported their age in years, sex assigned at birth, race, Hispanic or Latino/a/x ethnicity, and past-30-day vaping frequency. Age was treated as a continuous variable. Sex assigned at birth was assessed as male or female. Race was assessed using a multi-category self-identification item, and Hispanic or Latino/a/x ethnicity was assessed separately as yes/no. Past-30-day vaping frequency was assessed as the self-reported number of days participants used an e-cigarette or vaping device during the previous 30 days.

### Analytic Strategy

To address RQ1, we estimated cross-classified multilevel linear models (MLM) examining associations of cognitive engagement, social relevance, emotional arousal, and emotional valence with PME. All four perceptual dimensions were entered simultaneously as fixed effects, allowing their associations with PME to be estimated while adjusting for the other dimensions. Prior to estimation, ratings on each perceptual dimension were standardized (z-scored) within dimension across all message evaluations, so that coefficients reflect the change in PME associated with a 1-SD increase in a given dimension and could be compared across predictors; PME was retained on its original 1-5 metric. Because each participant evaluated multiple messages and each message was evaluated by multiple participants, models included crossed random intercepts for participant and message. Models additionally adjusted for age, sex assigned at birth, race, Hispanic ethnicity, and past-30-day vaping frequency.

We next used an Explainable Boosting Machine (EBM), implemented with the Explainable Boosting Regressor in the open-source *interpret* Python package,^28–30^ to probe potential nonlinear associations between the four perceptual dimensions and PME that may not be captured by the linear MLMs. EBM is an interpretable generalized additive modeling approach that uses boosting to estimate flexible shape functions for individual predictors while retaining predictor-level interpretability.^28,29^ For the EBM analyses, perceptual-dimension scores were retained on their original 1-9 scales because standardization was not required for model estimation and retaining the original metric facilitated interpretation of the learned shape functions. The 8,100 message evaluations were randomly divided into training (80%) and held-out test (20%) sets, and predictive performance was evaluated using root mean squared error (RMSE) and *R²* in the held-out test set. Learned shape functions were examined to characterize the form of the association between each perceptual dimension and predicted PME across its observed range.

To formally test the nonlinear valence pattern suggested by the EBM, we estimated piecewise (breakpoint) multilevel models with the same crossed random intercepts for participant and message. Valence was entered on its original 1-9 metric. We first compared linear and quadratic specifications using a likelihood-ratio test, then allowed separate valence slopes below and above a breakpoint, selecting the breakpoint that minimized model deviance across candidate values from 2 to 8. Segment-specific slopes were estimated without covariates and again adjusting for the other three perceptual dimensions and participant characteristics. To assess whether the valence pattern reflected overlap with emotional arousal, we examined valence-arousal correlations and variance inflation factors (VIFs) and re-estimated the adjusted model omitting arousal and, for comparison, omitting cognitive engagement and social relevance.

To address RQ2, we implemented a data-driven search for high-performing profiles, defined as combinations of perceptual characteristics associated with high predicted PME. Using the fitted EBM, we generated predicted PME for all unique combinations of cognitive engagement, social relevance, emotional arousal, and emotional valence observed across the 8,100 message evaluations (2,369 unique combinations). For each candidate profile, we calculated a PME-drop score as the difference between the maximum predicted PME and the profile’s predicted PME. Profiles with predicted PME within one held-out test-set RMSE of the maximum were retained as near-maximum predicted-PME profiles (i.e., high-performing profiles) for subsequent profile discovery. The retained 487 profiles were standardized before *k*-means clustering so that each perceptual dimension contributed comparably to the distance-based clustering solution. The standardized profiles were then grouped using k-means clustering.^31^ The number of clusters was selected by comparing average silhouette widths across candidate values of *k* (*k* = 2-29), with the five-cluster solution yielding the highest silhouette value (i.e., providing the strongest separation). Cluster centroids were then examined to characterize distinct combinations of cognitive, social, and emotional perceptions associated with high predicted PME.^32^ Analyses were conducted in 2026.

## Results

Across the 229-message corpus, PME averaged 3.67 (*SD* = 0.19). Mean message-level scores for the four perceptual dimensions were 6.11 (*SD* = 0.41) for cognitive engagement, 5.97 (*SD* = 0.39) for social relevance, 5.85 (*SD* = 0.41) for emotional arousal, and 5.66 (*SD* = 0.44) for emotional valence.

To address *RQ1*, cross-classified MLM models showed positive associations of cognitive engagement (*b* = 0.20, 95% CI [0.18, 0.22], *p* < .001), social relevance (*b* = 0.15, 95% CI [0.13, 0.17], *p* < .001), and emotional arousal (*b* = 0.06, 95% CI [0.04, 0.08], *p* < .001) with PME. Emotional valence showed no significant linear association with PME (*b* = 0.00, 95% CI [-0.02, 0.02], *p* = .868). Estimates were adjusted for participant demographics and past-30-day vaping frequency, with crossed random intercepts specified for participants and messages (see Table 2). Coefficients reflect the change in PME associated with a 1-SD increase in each perceptual dimension. Multicollinearity among the four dimensions was low (VIFs ranged from 1.06 to 1.63).

**Table 2.** Associations of the Four Perceptual Dimensions with PME.

| Dimensions | PME |  |  |
| --- | --- | --- | --- |
|  | *b* | 95% CI | *p* |
| Cognitive engagement | 0.20 | [0.18, 0.22] | <.001 |
| Social relevance | 0.15 | [0.13, 0.17] | <.001 |
| Emotional arousal | 0.06 | [0.04, 0.08] | <.001 |
| Emotional valence | 0.00 | [−0.02, 0.02] | .868 |

*Note.* Results are based on 8,100 message evaluations from 540 participants evaluating 229 messages. Coefficients (*b*) are fixed-effect estimates from a cross-classified multilevel model with crossed random intercepts for participants and messages, adjusting for age, sex assigned at birth, race, Hispanic ethnicity, and past-30-day vaping frequency. Perceptual dimensions were z-standardized and PME was retained on its original 1–5 scale, so each coefficient represents the change in PME associated with a 1-SD increase in the corresponding perceptual dimension.

The EBM further characterized potential nonlinear associations between the four perceptual dimensions and PME (Figure 1). Cognitive engagement, social relevance, and emotional arousal showed largely monotonic positive relationships with predicted PME. Emotional valence, in contrast, showed a nonlinear U-shaped pattern, with both more negative and more positive valence associated with higher predicted PME and more neutral valence associated with lower predicted PME. Predictive performance in the held-out test set was RMSE = 0.76 and *R²* = 0.35.

**Figure 1.**
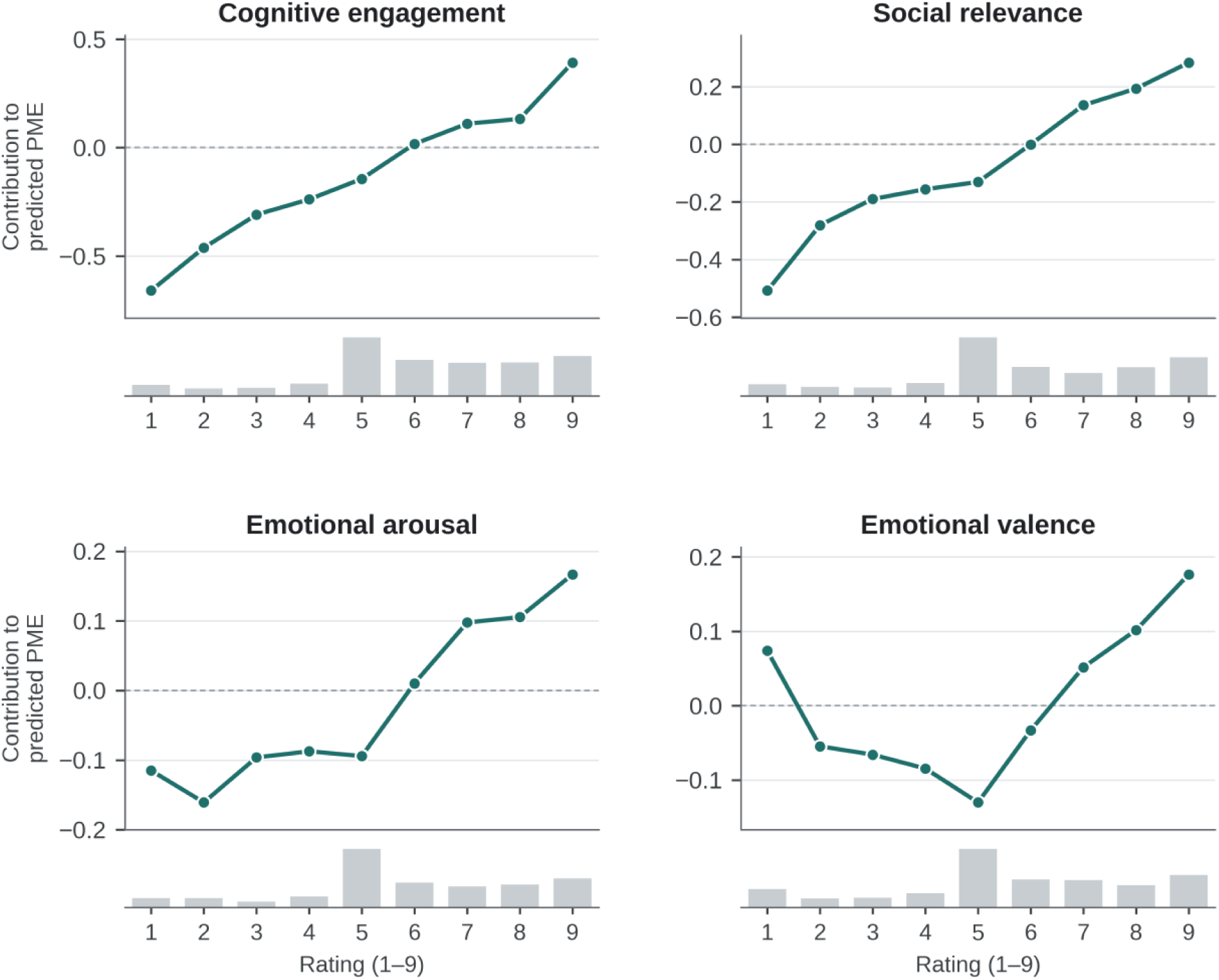
Main Effect of 4 Dimensions on PME from EBM. *Note*. Each panel plots the learned shape function for one perceptual dimension from the Explainable Boosting Machine. The x-axis is the message-evaluation rating on that dimension (1-9); the y-axis is that dimension’s additive contribution to predicted PME, expressed in PME units (1-5 scale). Gray bars beneath each panel show the distribution of ratings on that dimension across the 8,100 message evaluations.

Breakpoint analyses corroborated the nonlinear valence pattern. In a model with valence as the only predictor, the linear slope was near zero (*b* = 0.003, *p* = .555), whereas adding a quadratic term significantly improved fit (χ²(1) = 56.0, *p* < .001). Across candidate breakpoints, deviance was minimized at the scale midpoint of 5. In the resulting breakpoint model, PME increased as valence became more negative below the midpoint (*b* = −0.068 per scale point, *p* < .001) and as valence became more positive above it (*b* = 0.027, *p* = .013). After adjusting for the other three perceptual dimensions and participant characteristics, the negative-segment slope remained significant (*b* = −0.043, *p* < .001), whereas the positive-segment slope was attenuated and no longer significant (*b* = 0.015, *p* = .164); the breakpoint specification nonetheless continued to improve fit over a linear valence term (χ²(2) = 40.1, *p* < .001).

Collinearity did not account for these patterns. Valence and arousal were only weakly correlated overall (*r* = .17) and within the negative (*r* = −.24) and positive (*r* = .19) segments, and all VIFs in the adjusted main-effects model were below 1.2. Omitting arousal left the adjusted positive-segment slope essentially unchanged (*b* = 0.016, *p* = .131), whereas omitting cognitive engagement and social relevance largely restored it (*b* = 0.024, *p* = .028). The attenuation of the positive arm therefore reflected overlap with cognitive engagement and social relevance rather than with arousal.

To answer *RQ2*, 487 profiles met the prespecified criterion for high predicted PME. Clustering these profiles yielded five distinct high-PME profiles. The clusters reflected distinct combinations of cognitive engagement, social relevance, emotional arousal, and emotional valence, while predicted PME varied relatively little across clusters (4.10-4.25; Figure 2). For ease of interpretation, we assigned descriptive labels to the five clusters based on their relative patterns across the four perceptual dimensions. These labels are intended as interpretive shorthand and do not indicate objectively coded message features or appeal types. These included a *maximally engaging* profile characterized by uniformly elevated cognition, arousal, valence, and social relevance (Cognitive = 8.53; Arousal = 7.55; Valence = 7.29; Social = 7.73), and a *threat-based* profile that paired similarly high cognition, arousal, and social relevance with strongly negative valence (Cognitive = 8.05; Arousal = 7.21; Valence = 2.04; Social = 7.75). Three additional profiles produced comparably high predicted PME (4.10-4.16) via more differentiated configurations. For instance, the *calm rational* profile combined high cognitive engagement and social relevance with notably low arousal and moderately positive valence (Cognitive = 8.72; Arousal = 3.47; Valence = 6.76; Social = 7.88; PME = 4.16), the *intellectual depth* profile emphasized very high cognition and high arousal with only moderate valence but low social relevance (Cognitive = 9.00; Arousal = 7.00; Valence = 5.58; Social = 3.89; PME = 4.10), and the *positive affect* profile achieved similar PME with high arousal, positive valence, and high social relevance despite comparatively lower cognition (Cognitive = 6.34; Arousal = 7.64; Valence = 7.50; Social = 7.97; PME = 4.10).

**Figure 2.**
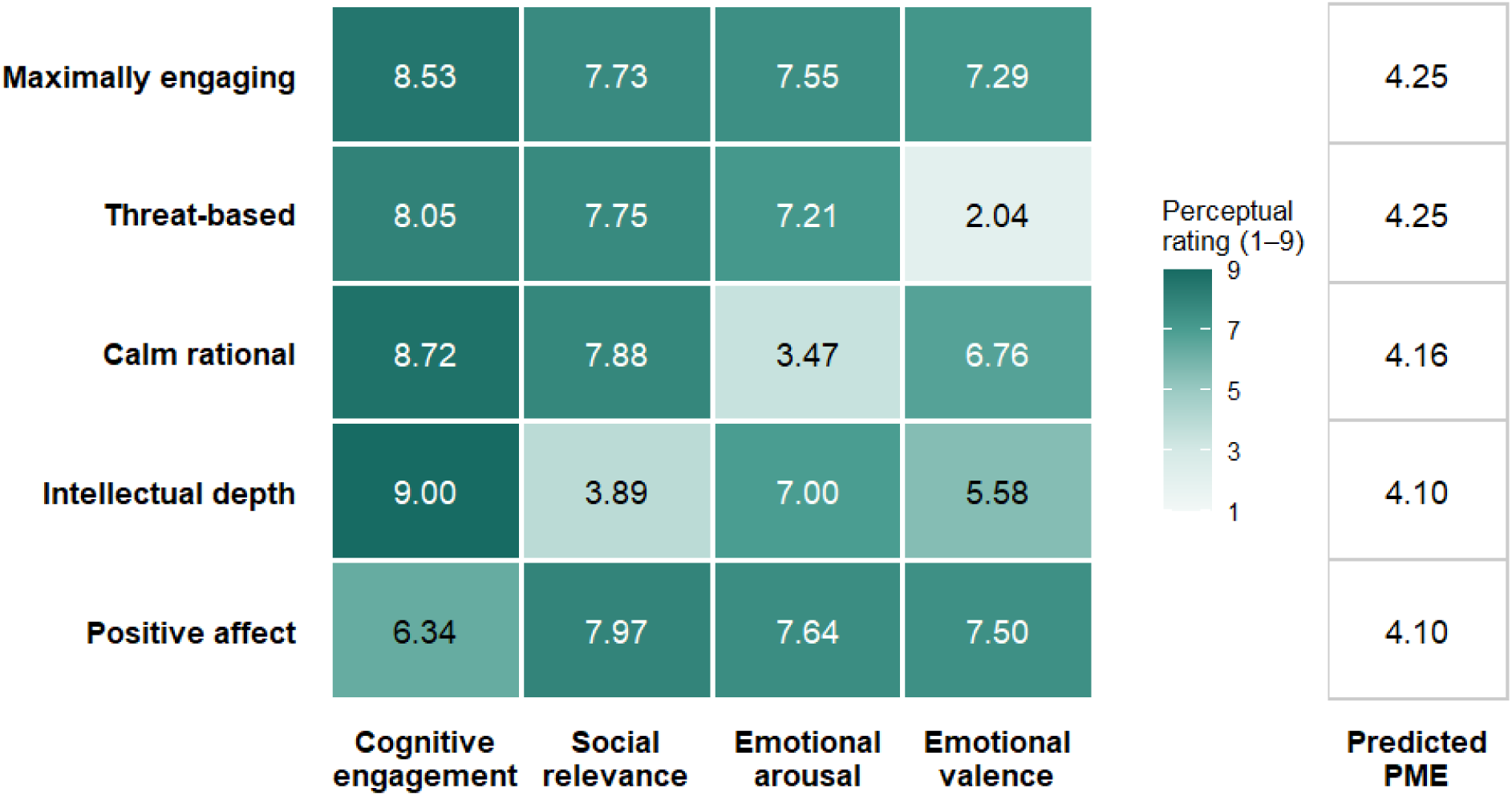
Five High-Predicted-PME Perceptual Profiles. *Note*. Cell values represent mean perceptual ratings within each cluster on the original 1–9 scales, with darker shading indicating higher ratings. Predicted PME represents the mean model-predicted PME across perceptual configurations assigned to each cluster. Cluster labels were assigned as descriptive shorthand based on the relative pattern of perceptual ratings within each cluster and should not be interpreted as objectively coded message features or appeal types.

In aggregate, while these profiles do suggest that cognitive engagement and perceived social relevance often co-occur in high-PME routes (Clusters 1, 2, 3, 5), persuasive success can also arise from alternative compensatory structures (e.g., cognitively intense, arousing “intellectual depth” messages with weaker social relevance), underscoring that persuasive success does not depend on a single ideal configuration of message features; instead, multiple distinct psychological profiles can yield similarly high levels of perceived effectiveness. Simply put, these findings highlight the importance of considering configurational, rather than purely additive, models of persuasion.

## Discussion

This study examined how cognitive, social, and emotional perceptions jointly characterize the perceived effectiveness of vaping public education messages among young adults who vape. Across a corpus of 229 messages, cognitive engagement and social relevance showed the strongest positive linear associations with PME, followed by a smaller positive association for emotional arousal. Emotional valence showed little linear association with PME but a pronounced nonlinear pattern in the EBM. The profile analysis further suggested that high predicted PME was not characterized by a single perceptual configuration, but by several distinct combinations of cognitive, social, and emotional responses.

The comparatively strong associations for cognitive engagement and social relevance suggest that how messages engage recipients’ thinking and connect vaping to socially meaningful consequences may be particularly important during formative message evaluation. The cognitive-engagement finding is consistent with persuasion theories that emphasize the role of elaborative processing in shaping responses to persuasive messages^19^. It is also consistent with vaping communication research showing that prevention messages can promote cognitive elaboration and that such elaboration may contribute to subsequent message effects.^16^ The social-relevance finding similarly complements evidence that social and normative considerations, including injunctive norms, can play an important role in responses to vaping prevention communication.^16,25^ Importantly, these dimensions were entered simultaneously in the MLMs, suggesting that each contributed information beyond the other perceptual characteristics assessed here. Emotional arousal was also positively associated with PME, although its comparatively smaller independent association suggests that arousal may provide less information about PME than cognitive engagement or social relevance.

The divergence between the linear and nonlinear analyses was especially informative for emotional valence. Valence showed essentially no linear association with PME, yet the EBM indicated higher predicted PME toward both the negative and positive ends of the valence continuum and lower predicted PME near the middle. Thus, the null linear coefficient alone would have obscured potentially meaningful heterogeneity in this association. This pattern suggests that strongly valenced messages may warrant consideration during formative testing, whether they evoke negative affect around vaping-related consequences or more positive responses related to autonomy or other desirable outcomes. However, because valence was not experimentally manipulated and the profile analysis was model-derived, these findings should not be interpreted as evidence that increasing emotional extremity will causally increase message effectiveness.

One potential concern is that the valence pattern reflects emotional intensity rather than valence per se, because strongly negative or positive messages may also be more arousing. Although valence and arousal are conceptually distinct dimensions of affective experience,^21^ they were modestly correlated in our data, particularly toward the extremes of the valence scale. However, collinearity diagnostics indicated little overlap, and removing arousal from the model did not change the valence slopes, suggesting that the U-shaped pattern is not an artifact of arousal. At the same time, the two arms of the pattern were not equally robust. The association with more negative valence held after adjusting for the other perceptual dimensions, whereas the association with more positive valence was weaker and was largely accounted for by cognitive engagement and social relevance. Positively valenced messages may therefore be most effective when they also prompt reflection or evoke socially meaningful consequences.

The profile analysis extends these dimension-specific findings by showing that similarly high predicted PME can arise from different combinations of perceptual responses. Several high-performing profiles combined strong cognitive engagement and social relevance but differed in arousal and valence, whereas other profiles achieved comparable predicted PME despite comparatively weaker scores on one dimension. These findings suggest a configurational view of formative message evaluation in which no single perceptual characteristic must be maximized in every message. Instead, different combinations of cognitive, social, and emotional responses may characterize messages with similarly high predicted PME. Importantly, these profiles were generated from the fitted model rather than experimentally manipulated message types and should therefore be viewed as candidate message “recipes” for subsequent validation rather than established design prescriptions.

These findings suggest that formative message testing may benefit from assessing multiple perceptual dimensions jointly rather than selecting messages on a single criterion. Cognitive engagement and social relevance emerged as particularly consistent correlates of PME. The nonlinear valence pattern was more robust at the negative end, whereas the positive-valence association was attenuated after accounting for cognitive engagement and social relevance. Thus, both negatively and positively valenced approaches may warrant consideration during formative testing, but positively valenced messages may be most promising when they also prompt reflection or evoke socially meaningful consequences. The identified profiles could therefore serve as candidate “recipes” for selecting or refining messages for subsequent experimental testing, rather than as prescriptive formulas for message design.

## Limitations

Several limitations should be considered. First, the primary outcome was PME rather than behavioral effectiveness. Although PME has demonstrated prospective associations with tobacco-related outcomes, including quit intentions and cessation behavior,^12^ the identified profiles require prospective validation against changes in beliefs, intentions, and vaping behavior. Second, the four perceptual dimensions and PME were assessed following the same message exposure, raising the possibility that shared-method variance or a more general evaluative response to the message contributed to their associations.^33^ Experimental studies that independently manipulate these dimensions would provide stronger evidence regarding their causal contributions. Third, cognitive engagement and social relevance were assessed using study-specific single-item measures to minimize burden during repeated message evaluation. Future work should further validate these measures and examine whether multi-item operationalizations yield similar patterns. Fourth, the high-PME profiles were generated from a fitted model for the combinations of ratings observed in the data and therefore represent model-implied perceptual configurations rather than profiles of individual messages in the corpus. Their robustness should be examined using alternative profile-selection thresholds, clustering specifications, and independent message samples. Relatedly, the training and held-out test sets for the Explainable Boosting Machine were formed by randomly splitting the 8,100 message evaluations rather than the 229 messages, so evaluations of the same message could appear in both sets; validation with message-level grouping would provide stronger evidence that the model generalizes to new messages. Finally, participants were recruited from an online panel of U.S. YAs who vape using demographic quotas, and the 229-message corpus was intentionally heterogeneous but does not represent the universe of vaping public education messages. Generalizability to adolescents, older adults, other tobacco-use populations, and other message sets remains to be established.

## Conclusions

Across a diverse corpus of vaping public education messages, cognitive engagement, social relevance, emotional arousal, and emotional valence showed distinct linear and nonlinear associations with perceived message effectiveness. High predicted PME was associated with multiple combinations of these perceptual characteristics rather than a single message profile. These findings suggest that multidimensional perceptual assessment may offer a useful framework for formative message testing and for identifying candidate message strategies for subsequent experimental and behavioral validation.

## Data Availability

De-identified, aggregated message-level ratings are available from the corresponding author upon reasonable request. The full stimulus corpus is not publicly redistributed because third-party copyright applies to a subset of the messages, which were adapted from existing campaign materials or online images.

## Data and Materials Availability

De-identified, aggregated message-level ratings are available from the corresponding author upon reasonable request. The full stimulus corpus is not publicly redistributed because third-party copyright applies to a subset of the messages, which were adapted from existing campaign materials or online images. Example messages are shown in Table 1.

## Ethics Approval

The study protocol was approved by the University of California, Santa Barbara Institutional Review Board (Protocol Number: 5-24-0806). All participants provided electronic informed consent before participation.

